# Seroprevalence of measles and varicella in healthcare workers in Chonburi province, Thailand between October 2022 and January 2023

**DOI:** 10.1101/2024.02.18.24303008

**Authors:** Apirat Katanyutanon, Wichai Thanasopon, Chaninan Sonthichai, Piyada Angsuwatcharakorn, Jira Chansaenroj, Ratchathorn Nakabut, Sarandhorn Naritpavalun, Yanathep Prasitsomsakul, Paranyu Surakhot, Phuvich Pompim, Nasamon Wanlapakorn, Yong Poovorawan

## Abstract

**Introduction:** Health care workers (HCWs) face the risk of contracting and transmitting vaccine-preventable infectious diseases (VPDs). Therefore, it is crucial to evaluate the immune status of HCWs against certain VPDs, such as measles and varicella zoster virus (VZV). This study aimed to determine age-specific seropositivity rates of anti-measles and anti-VZV IgG in HCWs working in Chonburi province, Thailand, and to develop a measles and varicella vaccination policy for Thai HCWs.

**Methods:** A total of 266 HCWs in Chonburi Province, Thailand, were enrolled in this study between October 2022 and January 2023. Participants were divided into six age groups: 21-30, 31-40, 41-50, 51-60, 61-70, and >70 years. Anti-measles and anti-VZV IgG levels were evaluated using commercial ELISA kits (EUROIMMUN, Lübeck, Germany).

**Results:** The overall seropositivity rates for measles and varicella were 85.0% and 81.2%, respectively. The lowest seropositivity rates for both measles and varicella were found among the 21-30-year-old group. Furthermore, seropositivity rates increased with age, reaching 100% among people over the age of 60 years.

**Conclusions:** To prevent measles and VZV outbreaks in HCWs, it is crucial to implement catch-up measles and varicella vaccination programs, particularly among HCWs younger than 30 years of age.

## Introduction

Measles and varicella zoster virus (VZV) are extremely contagious diseases that manifest with skin eruptions. The secondary attack rate of measles and varicella ranges between 70% and 90% among susceptible exposed individuals [1]. The basic reproduction number of measles and varicella ranges from 13 to 18 and 3.7 to 5.0, respectively [2, 3]. The measles vaccine was developed in the 1970s and was integrated into the Expanded Programme on Immunization (EPI) worldwide in 1974 [4]. The live attenuated varicella vaccine, derived from the VZV Oka strain, has been licensed for more than a decade [5, 6]. Despite the decline in the incidence of measles and varicella resulting from widespread vaccination efforts, outbreaks of these two diseases have been reported in settings where children gather, such as childcare centers and schools, in hospitals, institutionalized children and adults’ facilities, refugee camps, and among healthcare workers (HCWs) [7–10].

HCWs are at increased risk for measles than the general population. In the United States, 78 documented cases of measles were attributed to transmission within healthcare facilities [11]. Furthermore, 29 healthcare personnel contracted the virus through occupational exposure, with one individual transmitting the measles virus to a patient. Consequently, the CDC’s Advisory Committee on Immunization Practices (ACIP) recommends that HCWs who were born before 1957 or who lack serological evidence of immunity or prior vaccination should receive a two-dose regimen of the measles-mumps-rubella (MMR) vaccine, with a minimum interval of four weeks between doses [12].

The VZV vaccine, also known as chickenpox vaccine, is a live-attenuated vaccine for protection against chickenpox (varicella) [7, 13]. The currently licensed varicella vaccines are highly effective in preventing varicella and its complications [14, 13]. However, varicella outbreaks have been reported, including among healthcare workers in Thailand [15]. Previous seroprevalence studies showed high levels of seronegativity among healthcare workers ranging from 14–15.6% in Malaysia and Saudi Arabia, to as high as 51% among first-year medical and engineering students in Sri Lanka [16–18]. In Thailand, the varicella vaccine is an optional vaccine. Thus, the coverage is low. A previous study of VZV seroprevalence in Thailand conducted among university students found that 26% were seronegative to anti-VZV antibodies, suggesting that one in four young Thai adults remained susceptible to varicella [19]. The CDC’s ACIP recommends that for varicella, HCWs born after 1970, without a history of varicella and/or a negative result on the antibody screening test, are advised to receive a two-dose vaccination series.

To develop a measles and varicella vaccination policy for Thai HCWs, it is important to understand the seroprevalence of these two diseases. Currently, there are neither serological screening nor a catch-up vaccination campaign in the HCWs working in Chonburi Province, Thailand. This study aimed to investigate the immune status of HCWs against measles and varicella in a variety of healthcare settings located in Chonburi province, Thailand and to define an appropriate vaccination program among HCWs.

## Methods

### Study design and ethical considerations

This cross-sectional study was part of the large-scale serosurvey conducted in 11 districts in Chonburi province, Thailand, between October 2022 and January 2023 [20]. In the large-scale serosurvey, there was a total of 1193 general population and 266 HCWs. The cluster random sampling method was used to enroll participants. The stratified clusters within the 11 districts were divided into urban and rural strata. From each district, each cluster was selected using the probability proportionate-to-size sampling method. Healthcare workers were selected from distinct-level health facilities within each district using consecutive sampling in each district. In the present study, only serological samples from 266 HCWs were analyzed.

The protocol received approval from the Institutional Review Board of the Faculty of Medicine of Chulalongkorn University (IRB numbers 0706/65). This study adhered to the ethical principles stated in the Declaration of Helsinki and followed the principles of good clinical practice. Prior to their participation in the study, written consent was obtained from each participant.

### Participants and Sample Collection

HCWs were approached for eligibility. Exclusion criteria included aged <18 years or > 80 years, having immunosuppressive disorders, malignancy, or diseases that require immunosuppressive agents. A structured questionnaire was applied to voluntary HCWs. The data collected in this study included age, sex, history of measles and varicella infection, and previous vaccination.

Blood samples of 3-5 milliliters (mL) were collected and subsequently subjected to centrifugation to obtain serum samples, which were aliquoted and stored at −20°C until further laboratory tests.

### Laboratory testing

#### Seroprevalence of Measles and VZV

Commercial enzyme-linked immunosorbent assay (ELISA) kits were used according to the manufacturer’s instructions to measure IgG concentrations against measles and VZV (EURROIMMUN, Lubeck, Germany). Serum samples were initially diluted 1:100 and then further diluted to obtain values within the detection range. Anti-measles and anti-VZV IgG concentrations are expressed in international units per liter (IU/L). The cut-off for seropositivity and seroprotection rate against measles in this study was ≥ 275 IU/L. The cut-off for seropositivity and seroprotection rate against varicella in this study was ≥ 110 IU/L.

#### Statistical Analysis

Data on the seropositivity rates of anti-measles IgG and anti-VZV IgG are presented as numbers and percentages. Geometric mean titers (GMT) were calculated. The associations between sex and GMT were performed using Person’s chi-square test. Comparison of logarithmic-transformed GMTs between groups was compared using one-way ANOVA. All statistical analyses were performed with SPSS v23.0 (IBM Corp., Chicago, IL). Figures were generated using GraphPad Prism v9.4.1 (GraphPad Software, San Diego, CA). A *p*-value < 0.05 was considered statistically significant.

## Results

### Demographic characteristics of study participants

In this study, a total of 266 HCWs were included for the seroprevalence studies of measles and varicella. The mean age was 38.3 years (Table 1). There were more female than male participants in this study (77.8% female versus 22.2% male). Most of the HCWs were between 21-30 years old, followed by 31-40 years and 41-50 years old. Only 5.6% of the HCWs (15/266) indicated that they had contracted measles, while 21.8% (58/266) reported having been vaccinated against measles. On the contrary, up to 48.1% (128/266) indicated that they had contracted varicella, while only 4.9% (13/266) reported having been vaccinated against varicella.

**Table 1.** Demographic characteristics of the HCWs included in this study (N=266)

|  |  |
| --- | --- |
| **Mean age, years (SD)** | 38.3 (11.5) |
| **Sex** | N (%) |
| Male | 59 (22.2) |
| Female | 207 (77.8) |
| **Past history of measles** | N (%) |
| Yes | 15 (5.6) |
| No | 230 (86.5) |
| Uncertain | 21 (7.9) |
| **Previous vaccination against measles** | N (%) |
| Yes | 58 (21.8) |
| No | 75 (28.2) |
| Uncertain | 133 (50.0) |
| **Past history of varicella** | N (%) |
| Yes | 128 (48.1) |
| No | 122 (45.9) |
| Uncertain | 16 (6.0) |
| **Previous varicella vaccination** | N (%) |
| Yes | 13 (4.9) |
| No | 149 (56.0) |
| Uncertain | 104 (39.1) |

SD = standard deviation

### Seroprotection rates and GMTs against measles and VZV in healthcare workers

Table 2 shows the proportion of HCWs who achieved seroprotection rates against measles and their GMTs by age groups. Overall, 85% of the HCWs in this study had an anti-measles IgG of > 275 IU/L with a GMT of 1128.0 (964.5 – 1320.0) IU/L. A high seroprotection rate (>98%) was observed in older individuals 41 years of age. The lowest seroprotection rate was found among HCWs in the age group of 21–30 years (68.2%). HCWs in the 41-50- and 51-60-year-old age groups had significantly higher anti-measles IgG GMTs than the 21-30-old age group as shown in Table 2 (*p*-value < 0.001). There were no significant differences in GMT between male and female HCWs (*p*-value = 0.457).

**Table 2.** Seroprevalence of anti-measles IgG in HCWs by age group and sex.

| **Variables** | **No.** | **Proportion of seroprotected individuals, N (%)** | **GMT (IU/L) (95% CI)** | **p-value (One-way ANOVA)** |
| --- | --- | --- | --- | --- |
| **All** | 266 | 226 (85.0) | 1128.0 (964.5 – 1320.0) |  |
| **Age groups (years)** |  |  |  |  |
| **21-30** | 88 | 60 (68.2) | 614.8 (454.5 – 831.7) | Reference |
| **31-40** | 75 | 65 (86.7) | 1036.5 (783.1 – 1372.0) | 0.030 |
| **41-50** | 54 | 53 (98.1) | 2095.8 (1584.0 – 2773.0) | <0.0001* |
| **51-60** | 43 | 42 (97.7) | 1969.9 (1512.0 – 2567.0) | <0.0001* |
| **61-70** | 4 | 4 (100) | 2135.1 (319.1 – 14285.0) | 0.193 |
| **>70** | 2 | 2 (100) | 1037.4 (2.1 – 522915.0) | 0.976 |
| **Sex** |  |  |  |  |
| **Male** | 59 | 52 (88.1) | 1261 (935.7 – 1699.0) | 0.457 |
| **Female** | 207 | 174 (84.1) | 1093 (909.7 – 1313.0) |  |

GMT: Geometric mean titer, \* = statistically significant

Table 3 shows the proportion of HCWs who achieved the seroprotection rate against varicella and their GMTs by age groups. A total of 81.2% (216/266) of HCWs achieved a seroprotection rate against varicella, with a GMT of 394.0 (315.4-492.0) IU/L. Similar to measles, a high seroprotection rate (>90%) was observed in older individuals from 41 years of age. The lowest seroprotection rate for varicella was also found among HCWs in the 21– 30-year-old age group (64.8%). HCWs in the 41-50- and 51-60-year-old age groups had significantly higher anti-VZV IgG GMTs than the 21-30-old age group as shown in Table 3 (*p*-value < 0.05). No significant differences in the seroprotection rate were found between male and female HCWs (*p*-value = 0.508). In particular, the seroprotection rate against measles and VZV gradually increased with age and reached 100% in HCWs over 60 years of age.

**Table 3.** Seroprevalence of anti-VZV IgG in HCWs by age group and sex.

| Variables | No. | Proportion of seroprotected individuals, N (%) | GMT (IU/L) (95% CI) | *p*-value (One-way ANOVA) |
| --- | --- | --- | --- | --- |
| **All** | 266 | 216 (81.2) | 394.0 (315.4 – 492.0) |  |
| **Age groups (years)** |  |  |  |  |
| **21-30** | 88 | 57 (64.8) | 239.6 (148.6 – 386.2) | Reference |
| **31-40** | 75 | 63 (84.0) | 326.7 (213.8 – 499.1) | 0.774 |
| **41-50** | 54 | 50 (92.6) | 617.7 (436.3 – 874.4) | 0.012* |
| **51-60** | 43 | 40 (93.0) | 791.4 (553.3 – 1132.0) | 0.002* |
| **61-70** | 4 | 4 (100) | 767.1 (127.8 – 4605.0) | 0.659 |
| **>70** | 2 | 2 (100) | 616.3 (33.1 – 11461.0) | 0.947 |
| **Sex** |  |  |  |  |
| **Male** | 59 | 50 (84.7) | 453.3 (268.2 – 766.0) | 0.508 |
| **Female** | 207 | 166 (80.2) | 378.5 (296.1 – 483.9) |  |

GMT: Geometric mean titer, \* = statistically significant

### Correlation between serological results and self-reported medical history

Table 4 shows the correlation between the seroprotection rates against measles and VZV and the history of previous vaccination and infection. HCWs with a self-reported history of previous measles infection or vaccination had protective levels of anti-measles IgG in 86.7-89.7% of cases, but not significantly higher than in those who did not report previous infection or vaccination. On the contrary, a history of previous varicella was more frequent among those who were immune to varicella (*p*-value = 0.018) than among those who were susceptible. HCWs who reported having no previous vaccination against varicella were more likely to be immune (*p*-value < 0.001), suggesting that the positive serological results were due to natural infection. The positive predictive value of the history of varicella was 89.84%, where it was 86.67% for the history of measles (Table 5). The positive predictive value of the history of varicella vaccination was 76.92%, where it was 89.66% for the history of measles vaccination. The negative predictive value of the was low for both measles and varicella infection and vaccination in this study.

**Table 4.** The Correlation between the serological results of anti-measles anti-VZV IgG with the self-reported medical history.

| Variables | Total (N=266) | Immune (seroprotected) N (%) | Nonimmune (susceptible) N (%) | *p*-value |
| --- | --- | --- | --- | --- |
| **Measles** |  |  |  |  |
| **History of vaccination** |  |  |  | 0.064 |
| **Yes** | 58 | 52 (89.7) | 6 (10.3) |  |
| **No** | 75 | 67 (89.3) | 8 (10.7) |  |
| **Uncertain or N/A** | 133 | 107 (80.5) | 26 (19.5) |  |
| **History of measles** |  |  |  | 0.864 |
| **Yes** | 15 | 13 (86.7) | 2 (13.3) |  |
| **No** | 230 | 195 (84.8) | 35 (15.2) |  |
| **Uncertain or N/A** | 21 | 18 (85.7) | 3 (14.3) |  |
| **Varicella** |  |  |  |  |
| **History of vaccination** |  |  |  | <0.001* |
| **Yes** | 13 | 10 (76.9) | 3 (23.1) |  |
| **No** | 149 | 132 (88.6) | 17 (11.4) |  |
| **Uncertain or N/A** | 104 | 74 (71.2) | 30 (28.8) |  |
| **History of varicella** |  |  |  | 0.018* |
| **Yes** | 128 | 115 (89.8) | 13 (10.2) |  |
| **No** | 122 | 89 (73.0) | 33 (27.0) |  |
| **Uncertain or N/A** | 16 | 12 (75.0) | 4 (25.0) |  |

N/A: Data not available.

**Table 5.** Positive predictive values and negative predictive values of the self-reported history of infection or vaccination.

|  |  | Positive predictive value (%) | Negative predictive value (%) |
| --- | --- | --- | --- |
| Measles | History of measles | 86.67 | 15.22 |
|  | History of vaccination | 89.66 | 10.67 |
| Varicella | History of varicella | 89.84 | 27.05 |
|  | History of vaccination | 76.92 | 11.41 |

## Discussion

Vaccination for HCWs is an essential step in protecting not only HCWs but also patients and communities [21, 22]. To establish an effective immunization program, a serological survey is necessary to obtain the immune status of HCWs, together with a history of previous infection and vaccination, are necessary. Our study showed high seroprotection rates of more than >90% for measles (anti-measles IgG > 275 IU/L) and VZV (anti-VZV IgG > 110 IU/L) in HCWs older than 41 years working in Chonburi Province, Thailand, even when the policy for the screening and vaccination campaign in HCWs has not been implemented. This preexisting immunity is likely to result from natural exposure before the EPI for measles and varicella vaccinations were in place for all infants. On the contrary, individuals between 21-30 years of age who were born under the universal measles vaccination for all infants at 9 months of age had the lowest seroprotection rate for measles. A recent seroprevalence survey in Thai adolescents also showed that, despite 70% documented one- or two-dose measles vaccination during childhood, only half of the adolescents had anti-measles IgG greater than 275 IU/L, likely due to the decline of antibodies without natural exposure or primary vaccine failure [23]. Thus, in this study, HCWs younger than 30 years of age who could have been vaccinated against measles under the EPI program could possibly have anti-measles immunity that waned overtime. Furthermore, the second dose of measles-containing vaccine coverage among Thai children reached more than 80% of the population only after 2016, suggesting that a large number of populations may have received only one dose of measles-containing vaccine during childhood [24]. Thus, it is important to prioritize a catch-up campaign for measles vaccination for young healthcare workers who had no history of two-dose measles immunization or had negative serology.

In the present study, the overall seroprotection rate for measles in HCWs was lower than in the general population over 26 years of age in our previous serosurvey conducted in 2014 (85% versus 97.7%) [25]. The younger generation of the 2030s, having been born under the Expanded Program on Immunization (EPI) with improved vaccine coverage, may possess vaccine-induced immunity that diminishes over time, resulting in lower seroprevalence rates during adulthood compared to the previous survey 10 years ago. Additionally, comparison of anti-measles seroprevalence among HCWs from different regions showed that HCWs from high-income countries had a higher rate of measles seroprevalence ranging from 87% to 97% than HCWs in our study [9].

Our study showed that the overall seroprotection rate against varicella was 81.2% This rate was lower compared to HCWs in Japan (96.3%) [26], Finland (99%) [27], Denmark (98.6%) [28], and Korea (91%) [29, 30]. In Thailand, older individuals acquired immunity to varicella through natural exposure, while younger individuals may have received varicella vaccination, which are optional vaccines, or may have contracted the disease during childhood. Therefore, younger HCWs are likely to have a low seroprotection rate compared to older HCWs due to the low coverage of optional vaccines and the lack of exposure to natural infection. Other countries also reported low anti-VZV seroprevalence rates in the young population [31]. Based on our results, it is recommended to prioritize varicella vaccination for HCWs in the 21-30 age group with negative serology and no record of two-dose varicella vaccination.

The correlation between self-reported history and seroprotected status showed that neither history of measles vaccination nor history of measles were associated with seroprotected status. Our results are consistent with other studies showing that self-reported vaccination history may not be predictive and reliable [27]. Therefore, when a measles vaccination record was not available, serologic tests should be performed to provide evidence of measles immunity. Unlike measles, we found that a history of previous varicella was more frequent among those who were immune to varicella. This is similar to the report of a Thai university student showing the significant association between the history of varicella and the seropositivity rate [19]. Consistent with a previous report, negative predictive values for both previous vaccination and infection were very low for both measles and varicella [30]. Our research indicates the necessity of implementing a universal record system to document vaccine administration for every individual in Thailand. The universal record system could potentially reduce unnecessary serologic screening and catch-up vaccinations.

This study has some limitations. First, the participants’ history of vaccination and previous infection was only obtained through questionnaires, which resulted in missing information and uncertain information regarding diagnosis, actual vaccination doses, and dates. Selecting HCWs from one province may not fully represent all HCWs in Thailand. A larger study that explores other regions of Thailand could provide a more comprehensive representation of the entire country. Additionally, the small sample size of participants older than 60 years makes it challenging to draw conclusive results.

## Conclusions

This study provides comprehensive information to inform policy makers and establish effective measles and varicella immunization programs for HCWs throughout the country. To prevent measles and VZV outbreaks in HCWs, it is crucial to conduct serological screening and implement catch-up measles and varicella vaccination programs, particularly among HCWs younger than 30 years of age.

## Acknowledgements

We greatly appreciate the kind contributions and collaboration of all participants. With all of their help, the interesting information obtained from this study could be used for the future development of vaccine strategies. We thank all staff from the hospitals in Chonburi Province for enrolling participants and staff from the Center of Excellence in Clinical Virology, Faculty of Medicine, Chulalongkorn University, for their help with the laboratory testing.

## Ethics approval

The study protocol was reviewed and approved by the Institutional Review Board (IRB), Faculty of Medicine, Chulalongkorn University (IRB 0706/65). Written informed consent was waived by the IRB.

## Conflict of interest statement

The authors have no conflict of interest to declare.

## Funding

This research was financially supported by the Health Systems Research Institute (HSRI), the National Research Council of Thailand (NRCT), the Research Chair Grant from the National Science and Technology Development Agency (P-15-50004), the Center of Excellence in Clinical Virology, Chulalongkorn University, and King Chulalongkorn Memorial Hospital and the Second Century Fund (C2F) of Jira Chansaenroj, Chulalongkorn University. The funders had no role in study design, data collection, analysis, publication decision, or manuscript preparation.

## Author contributions

Conceptualization: Y.P.; Data curation: A.K., W.T., J.C., N.W.; Formal analysis: A.K., J.C., N.W.; Funding acquisition: J.C., Y.P.; Investigation: A.K., W.T.; Methodology: A.K., J.C.; Project administration: J.C., R.N., S.N., Y.P., P.S., P.P.; Resources: A.K., W.T., C.S., P.A.; Software: J.C., N.W.; Supervision: Y.P.; Validation: A.K., N.W., Y.P.; Visualization: J.C.; Roles/Writing - original draft: A.K., J.C., R.N., S.N., Y.P., P.S., P.P., N.W.; and Writing – review & editing: A.K., N.W.

## Data availability statement

The authors confirm that all data supporting the findings of this study are available within the article.

## Notes

### Competing Interest Statement

The authors have declared no competing interest.

### Author Declarations

Institutional Review Board (IRB), Faculty of Medicine, Chulalongkorn University (IRB No.0706/65) and The Institutional Review Board of Chonburi Provincial Public Health Office, Ministry of Public Health (IRB No. 0024-2565), Thailand gave ethical approval for this work.

